# Dichotomy between the humoral and cellular responses elicited by mRNA and adenoviral vector vaccines against SARS-CoV-2

**DOI:** 10.1101/2021.09.17.21263528

**Authors:** Rahul Ukey, Natalie Bruiners, Hridesh Mishra, Pankaj K. Mishra, Deborah McCloskey, Alberta Onyuka, Fei Chen, Abraham Pinter, Daniela Weiskopf, Alessandro Sette, Jason Roy, Sunanda Gaur, Maria Laura Gennaro

## Abstract

Protection from severe disease and hospitalization by SARS-CoV-2 vaccination has been amply demonstrated by real-world data. However, the rapidly evolving pandemic raises new concerns. One pertains efficacy of adenoviral vector-based vaccines, particularly the single-dose Ad26.COV2.S, relative to mRNA vaccines. We investigated the immunogenicity of Ad26.COV2.S and mRNA vaccines in 33 subjects vaccinated with either vaccine class five months earlier on average. After controlling for time since vaccination, Spike-binding antibody and neutralizing antibody levels were higher in the mRNA-vaccinated subjects, while no significant differences in antigen-specific B cell and T cell responses were observed between the two groups. Thus, a dichotomy exists between humoral and cellular responses elicited by the two vaccine classes. Our results have implications for the need of booster doses in vaccinated subjects and might explain the dichotomy reported between the waning protection from symptomatic infection by SARS-CoV-2 vaccination and its persisting efficacy in preventing hospitalization and death.

---

With the COVID-19 pandemic still raging and new SARS-CoV-2 variants, such as Delta (B.1.617.2), exhibiting increased transmissibility ^1^, concerns have been raised about the efficacy of current vaccines in general as well as relative to each other. The SARS-CoV-2 vaccines that have received full approval or emergency use authorization by the US Food and Drug administration include the mRNA vaccines BNT162b2 (BioNTech-Pfizer) ^2^ and mRNA-1273 (Moderna) ^3^, which are administered in two doses, and the single-dose, adenoviral vector vaccine Ad26.COV2.S (Johnson and Johnson-Janssen) ^4^. Comparisons of protective immune responses elicited by these vaccines have focused on neutralizing titers in plasma [for example, ^5,6^]. Virus neutralization by plasma is critical to protect against viral infection, but understanding the efficacy and durability of vaccine-induced responses requires assessments of both humoral and cellular adaptive immune responses elicited by vaccination.

Here we used quantitative assays to compare antibody binding and neutralizing titers, antigen-specific B cell frequencies, and antigen-specific T cell responses in thirty-three participants with no history of SARS-CoV-2 infection, similarly divided between subjects having received mRNA vaccines (n = 16) or the adenoviral vector vaccine (n = 17). When we compared the two groups by age, gender, and co-morbidities, we found no difference in these variables except that for time elapsed since vaccination, which differed between the two groups (**Table 1**). Thus, as needed, the results of the immunological assays were adjusted by the time (in days) between full vaccine administration and blood collection for the study using linear regression. All methods are described in the *Supplement*.

**Table 1:** Demographics and clinical information of study participants, stratified by vaccine type.

|  | **Overall (n=33)** | **J&J (n=17)** | **mRNA (n=16)** | **p** |
| --- | --- | --- | --- | --- |
| **Age (years)** | 49.8 ± 15.6 | 52.3 ± 13.3 | 47.3 ± 17.7 | 0.066 |
| **Gender** |  |  |  |  |
| **Female** | 17/33<br>(51%) | 8/17<br>(47%) | 9/16<br>(56%) | 0.279 |
| **Male** | 16/33<br>(49%) | 9/17<br>(53%) | 7/16<br>(44%) |  |
| **BMI (kg/m<sup>2</sup>)** | 26.7 ± 5.3 | 27.4 ± 5.2 | 25.9 ± 5.4 | 0.413 |
| **Race** |  |  |  |  |
| **Not Hispanic or Latino** | 33/33<br>(100%) | 17/17<br>(100%) | 16/16<br>(100%) | - |
| **Ethnicity** |  |  |  |  |
| **Asian** | 10/33<br>(30%) | 4/17<br>(24%) | 6/16<br>(38%) | 0.350 |
| **Others** | 1/33<br>(3%) | 0/17<br>(0%) | 1/16<br>(6%) |  |
| **White** | 22/33<br>(67%) | 13/17<br>(76%) | 9/16<br>(56%) |  |
| **Comorbidities** |  |  |  |  |
| **Yes** | 10/33<br>(30%) | 5*/17<br>(29%) | 5*/16<br>(31%) | 0.909 |
| **No** | 23/33<br>(70%) | 12/17<br>(71%) | 11/16<br>(69%) |  |
| **Immunodeficiencies** |  |  |  |  |
| **Yes** | 3/33<br>(9%) | 0/17<br>(0%) | 3**/16<br>(19%) | 0.061 |
| **No** | 30/33<br>(91%) | 17/17<br>(100%) | 13/16<br>(81%) |  |
| **Days since vaccination** | 168.2 ± 57.9 | 189.7 ± 62.8 | 145.2 ± 43.3 | 0.025 |

Data are presented as mean ± standard deviation or proportion (n/N %). BMI, body mass index.
\*hypertension (n=6), obesity (n=3), diabetes (n=2), asthma (n=2), coronary artery disease (n=1) (some conditions were concurrent).
\*\*neutropenia (n=1), rheumatoid arthritis (n=1), use of corticosteroids (n=1).

## Antibody binding and neutralization

All vaccines express the full-length SARS-CoV-2 Spike protein ^2-4^. We analyzed plasma of all subjects for antibodies binding the receptor binding domain (RBD) of the S1 subunit of the SARS-CoV-2 Spike protein and for neutralizing antibodies. We chose RBD as target antigen of the antibody response, because the neutralizing activity of plasma is largely directed against RBD, as shown by us and others ^7-9^. The virus neutralization activity of plasma was measured with an assay utilizing replication-competent SARS-CoV-2 virus. We found that both Ab binding and neutralizing titers were higher in the mRNA-vaccinated group relative to adenoviral vector vaccinees (**Fig. 1AB**). The differences between groups were statistically significant after adjusting for days since vaccination (**Table 2**).

**Fig. 1.**
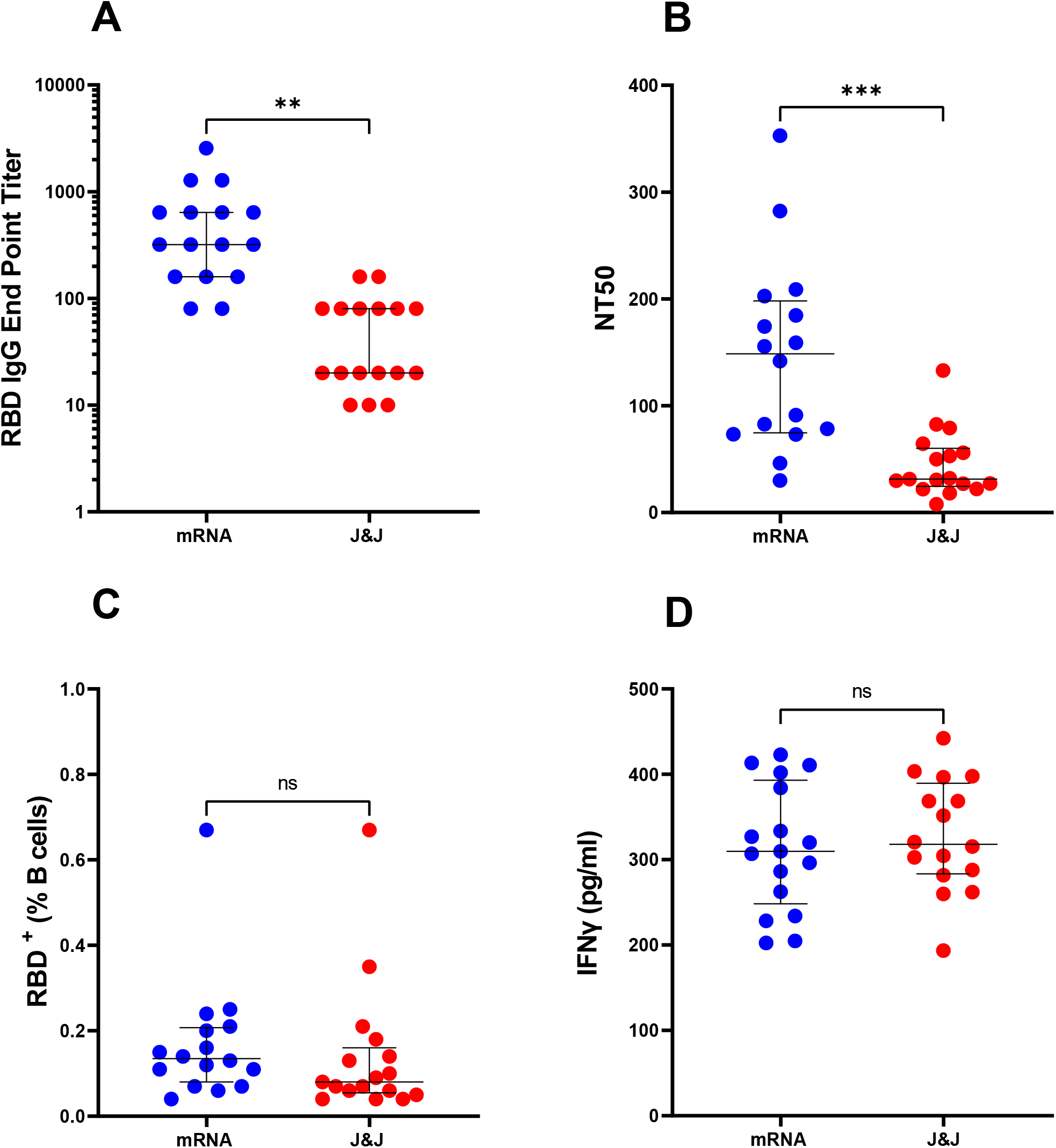
Humoral and cellular responses elicited by mRNA and adenoviral vector based COVID-19 vaccines. Each circle indicates one subject. Blue circles represent subjects who received two doses of mRNA vaccine (n=16) and red circles represent subjects who received the adenoviral vector-based (J&J) vaccine (n=17). The dot plots show (**A**) anti-RBD IgG antibody levels, (**B**) Neutralizing titers expressed as NT50 (reciprocal dilution of plasma yielding 50% neutralization of live SARS-CoV-2 virus), (**C**) Frequency (%) of RBD-specific B cells. B cells (CD19^+^CD 20^low^) were analyzed for RBD specificity utilizing dual fluorescent labelling of RBD tetramers. (**D**) IFNγ (pg/ml) production by antigen specific T cells. PBMCs from each subject were stimulated with a megapool of synthetic overlapping peptides covering the entire Spike (S) antigen. Supernatants were collected after 24 hours of stimulation. In all panels, the solid black lines represent the median and interquartile range. Statistical analyses were conducted by Mann-Whitney U test for unpaired samples (**, *p*≤0.01; ***, *p*≤0.001; ns, non-significant, *p*>0.05).

**Table 2:**
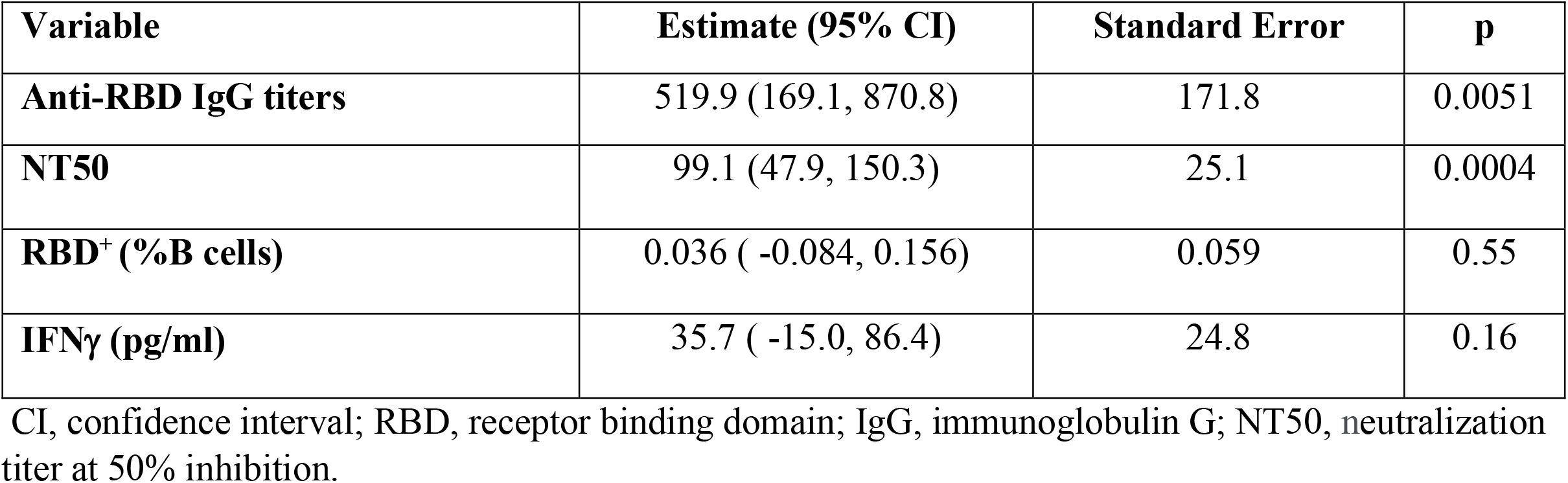
Estimates of mean difference in each measurement between mRNA and J&J (reference) vaccinees from a linear regression, adjusted for time since vaccination.

| **Variable** | **Estimate (95% CI)** | **Standard Error** | **p** |
| --- | --- | --- | --- |
| **Anti-RBD IgG titers** | 519.9 (169.1, 870.8) | 171.8 | 0.0051 |
| **NT50** | 99.1 (47.9, 150.3) | 25.1 | 0.0004 |
| **RBD<sup>+</sup> (%B cells)** | 0.036 ( -0.084, 0.156) | 0.059 | 0.55 |
| **IFNγ (pg/ml)** | 35.7 ( -15.0, 86.4) | 24.8 | 0.16 |

CI, confidence interval; RBD, receptor binding domain; IgG, immunoglobulin G; NT50, neutralization titer at 50% inhibition.

## RBD+ B cell frequencies

The levels of specific antibodies in the circulation physiologically decrease with time elapsed since exposure to antigen ^7,10,11^. Thus, assessing durability of vaccine-induced responses and protection from occurrence and clinical severity of breakthrough infection requires the evaluation of antigen-specific B cells and T cells. To analyze the memory B cell response elicited by the vaccine, we enumerated RBD-specific B cells (RBD-tetramer-positive CD19^+^CD20^low^) by multi-color flow cytometry (the gating strategy is shown in **Fig. S1**). We found that the differences in RBD-specific B cell frequencies between the mRNA- and adenoviral vector-vaccinated subjects were not statistically significant (**Fig. 1C and Table 2**).

## Interferon gamma (IFNγ) release assay

A straightforward method to assess antigen-specific T cells is measuring the production of T cell cytokines, such as IFNγ, by peripheral blood mononuclear cells (PBMC) stimulated ex vivo with peptides representing T cell epitopes, as performed for many infectious and non-infectious conditions (for example ^12^). We used a previously described peptide pool containing hundreds of overlapping 15-mers derived from the Spike protein (CD4_S) ^13^ for PBMC stimulation and detection of IFNγ release by ELISA. This assay showed no significant differences between the two groups of vaccinees (**Fig. 1D and Table 2**).

In conclusion, mRNA vaccination results in higher levels of circulating binding and neutralizing antibodies than the adenoviral vector counterpart (at least in the timeframe of our study, i.e., 5 months after vaccination on average), while the antigen-specific cellular responses to the two vaccine classes show no significant differences. Thus, a dichotomy exists between humoral and cellular immune responses elicited by the SARS-CoV-2 vaccines. One may postulate that the humoral response provides a “ready-to-go” response to reinfection that rapidly limits viral replication and the consequent development of symptoms. In contrast, memory immune responses, which require longer to be expressed even in vaccinated individuals, may best explain the persisting vaccine-induced protection against severe disease and hospitalization. Thus, the humoral vs cellular dichotomy seen in the present study might reflect that reported between the waning protection from symptomatic infection afforded by SARS-CoV-2 vaccination and its persisting efficacy in preventing hospitalization and death [^14^ and https://papers.ssrn.com/sol3/papers.cfm?abstract_id=3909743]. This scenario has implications for the expected effects of booster doses, which might counter the progressively increasing vulnerability to symptomatic infection by eliciting a vigorous antibody response while exerting no additional benefits on hospitalization and death rates among vaccinated, immunocompetent subjects. Future investigations will test this proposed scenario. An additional consideration is that, since vaccine-induced neutralizing antibodies are highly correlated with immune protection from symptomatic infection ^15,16^ – a correlation supported by recent murine studies ^17^, our data suggest that a booster dose of the Ad26.COV2.S is advisable, particularly in the face of the global rise of COVID-19 morbidity due to the highly infectious SARS-CoV-2 Delta variant ^1^. Indeed, the administration of a second vaccine dose of Ad26.COV2.S predictably induces a stronger antibody response than the primary vaccination, as per interim data by the manufacturer ^18^.

## Data Availability

The data that support the findings of this study are available on request from the corresponding author.

## Acknowledgements

We thank the 33 study participants that prompted this study; Dennis Burton and Pei-Yong Shi for providing biological reagents; and Martin Blaser and Karl Drlica for critical reading of the manuscript. This work was funded by NIH grants R01 HL149450, R01 HL149450-S1, U01 AI122285-S1, and UL1 TR003017, and NIH contract 75N9301900065.

## Materials and Methods

### Study participants

Thirty-three subjects who received either mRNA vaccines (n=16) or the adenoviral vector vaccine Ad26.COV2.S (n=17) were enrolled on August 9-10, 2021 at the Rutgers Robert Wood Johnson Medical School, New Brunswick, New Jersey, USA. All participants self-reported no history of SARS-CoV-2 infection and date of vaccination and consented to blood draws as well as collection of demographic data. All study activities were approved by the Rutgers Institutional Review Board (Pro2020000655).

### Biosafety protocols

All work involving blood products from SARS-CoV-2-infected subjects were performed in a biosafety level 2+ (BSL-2+) laboratory utilizing protocols approved by the Rutgers Institutional Biosafety Committee. All plasma samples were heat-inactivated at 56°C for 60 min before testing. Work involving live SARS-CoV-2 were performed in a biosafety level 3 (BSL-3) laboratory utilizing protocols approved by the Rutgers Institutional Biosafety Committee.

### Antibody binding by enzyme-linked immunosorbent assay (ELISA)

Antibody binding was performed by ELISA platform utilizing SARS-CoV-2 receptor binding domain (RBD) of the Spike protein as solid-phase antigen and standard operating procedures, as described ^1^. Each sample was tested in duplicate. End-point titers were calculated using an established cut-off ^1^ and background-subtracted data.

### Cell lines

Vero E6 were obtained from the American Type Culture Collection (ATCC), Manassas, USA; HeLa cells stably expressing ACE2 (HeLa-ACE2) were obtained from Dennis Burton at the Scripps Research Institute ^2^. All cell lines were maintained in high-glucose Dulbecco’s modified Eagle’s medium (DMEM; Corning, Manassas, USA) supplemented with 10% fetal bovine serum (FBS; Seradigm, Radnor, USA), 2mM L-glutamine, and 1% penicillin/streptomycin (Corning, Manassas, USA), and incubated in humidified atmospheric air containing 5% CO_2_ at 37°C.

### SARS-CoV-2 virus

The virus stock of mNeonGreen (mNG) SARS-CoV-2 was obtained from Pei-Yong Shi at the University of Texas Medical Branch at Galveston. The virus stock was produced using the virus isolate of the first patient diagnosed in the USA, in which the ORF7 of the viral genome was replaced with the reported mNG gene ^3^. Propagation of viral stocks was performed with Vero E6 cells using 2% FBS. The virus titers were determined by standard plaque assay utilizing Vero E6 cells ^4^ and recorded as plaque forming units per milliliter (PFU/mL).

### SARS-CoV-2 neutralization assay

HeLa-Ace2 cells were seeded in 96-well black optical-bottom plates at a density of 1 × 10^4^ cells/well in FluoroBrite DMEM (Thermo Fisher Scientific, Waltham, USA) containing 4% FBS (Seradigm), 2mM L-glutamine, and 1% penicillin/streptomycin (Corning, Manassas, USA), and incubated overnight at 37°C with 5% CO_2_. On the following day, each sample was subjected to two-fold serial dilution in DMEM without FBS, and incubated with mNG SARS-CoV-2 at 37°C for 1.5 hrs. The virus-plasma mixture was transferred to 96-well plates containing Hela-Ace2 cells at a final multiplicity of infection (MOI) of 0.25 (viral PFU:cell). For each sample, the starting dilution was 1:20 and the final dilution of 1:10,240. After incubating infected cells at 37°C for 20 hrs, mNG SARS-CoV-2 fluorescence was measured using a Cytation™ 5 reader (BioTek, Winooski, USA). Each sample was tested in duplicate. Relative fluorescent units were converted to percent neutralization by normalizing the sample-treatment to non-sample-treatment controls and plotting the data with a nonlinear regression curve fit to determine the titer neutralizing 50% of SARS-CoV-2 fluorescence (NT_50_).

### PBMC isolation and storage

Peripheral blood mononuclear cells (PBMC) were isolated by Ficoll density gradient centrifugation (Ficoll-Paque, GE Healthcare, Uppsala, Sweden), as described ^5^, cryopreserved in liquid nitrogen in FBS containing 10% dimethyl sulfoxide (DMSO; Thermo Fischer Scientific, Waltham, MA, USA), and stored until use.

### RBD-specific B cell immunostaining

To form RBD tetramers, biotinylated RBD (BioLegend, San Diego, CA, USA) was mixed in separate tubes with streptavidin conjugated with Alexa Fluor 647 or BV421 (BD Biosciences, San Jose, CA, USA) at a 4:1 molar ratio for 1 hour at 4°C. PBMCs were stained with fixable viability stain 780 (BD Biosciences, Franklin Lakes, USA), incubated for 10 minutes with human Fc receptor blocking reagent (BD Biosciences, Franklin Lakes, NJ, USA), and then stained with the two fluorescent RBD tetramers, antibodies to CD19-BV700 (HIB19, Bio Legend, San Diego, CA, USA) and CD20-PE-CF594 (2H7, BD Biosciences, San Jose, CA, USA) to detect RBD-specific B cells, and APC-Cy7-labelled antibodies against CD3 (UCHT1), CD4 (OKT4), CD14 (C1D3), and CD16 (CD16) (all from Thermo Fisher Scientific, Waltham, MA, USA) to eliminate non-B cells. Samples were incubated for 30 minutes on ice in the dark to allow for antibody binding, washed twice with FACS buffer (2 percent FBS in PBS), fixed for 20 minutes with 4 percent paraformaldehyde (PFA; Thermo Fisher Scientific, Waltham, MA, USA), and stored at 4°C overnight. At least 500,000 events were collected per sample utilizing a Fortessa X-20 flow cytometer (BD Biosciences, San Jose, CA, USA). After removing dead and non-B cells, B cells were separated as CD19^+^CD20^Lo^, and the frequency of B cells positive for both RBD tetramers was determined by using Flow Jo software (Flow Jo LLC, USA).

### SARS-CoV-2-derived peptides

A megapool (MP_S) of 253 15-mer synthetic peptides overlapping by 10-residues that cover the entire spike (S) antigen was generated based on predicted SARS-CoV-2 epitopes, as previously reported ^6,7^.

### IFNγ release assay

PBMC were washed in pre-warmed RPMI 1640 supplemented with 2 mM L-glutamine, 10% FBS, 100 U/ml penicillin, and 100 μg/ml streptomycin (complete RPMI) (all from Corning cellgro, Manassas, VA, USA), seeded in a 48-well cell culture plate at a density of 1 × 10^4^ cells/well in complete RPMI, and stimulated with 1μg/ml of the SARS-CoV-2 MP_S peptide pool or 0.1% DMSO (vehicle control). Cell culture plates were incubated for 24 hrs at 37°C in a 5% CO_2_ humidified atmosphere. Each sample was tested in duplicate. As positive control, two wells per sample were treated with a mixture of 25 ng/ml phorbol 12-myristate 13-acetate (PMA) (Sigma-Aldrich, St. Louis, MO, USA) and 0.5 μM ionomycin calcium salt (Enzo Life Sciences, Farmingdale, CT, USA) for 2 hrs. Supernatants were collected and levels of IFNγ in supernatants were assayed using a commercial Human IFNγ ELISA kit (BD Biosciences, San Jose, CA), according to manufacturer’s instructions.

### Statistical Analysis

Baseline demographic and other variables were tested using two-sample proportion test and Student’s t-test. All flow cytometry data were analyzed with FlowJo v12 software (FlowJo LLC, Ashland, OR, USA). Measurements from all immunological assays were compared between the two study groups using the Mann-Whitney U test. Linear regression was performed to assess the dependency of immunological measurements on type of vaccination while adjusting for time (days) elapsed since vaccination. Correlation was analyzed using the Spearman’s rank correlation. For all tests, *p*<0.05 was considered significant. Statistical analyses were performed with Stata (version 17, StataCorp LLC, College Station, USA) and GraphPad Prism version 8.4 (Graph Pad Software Inc., La Jolla, USA).

## Supplemental Figure Legend

**Fig. S1.**
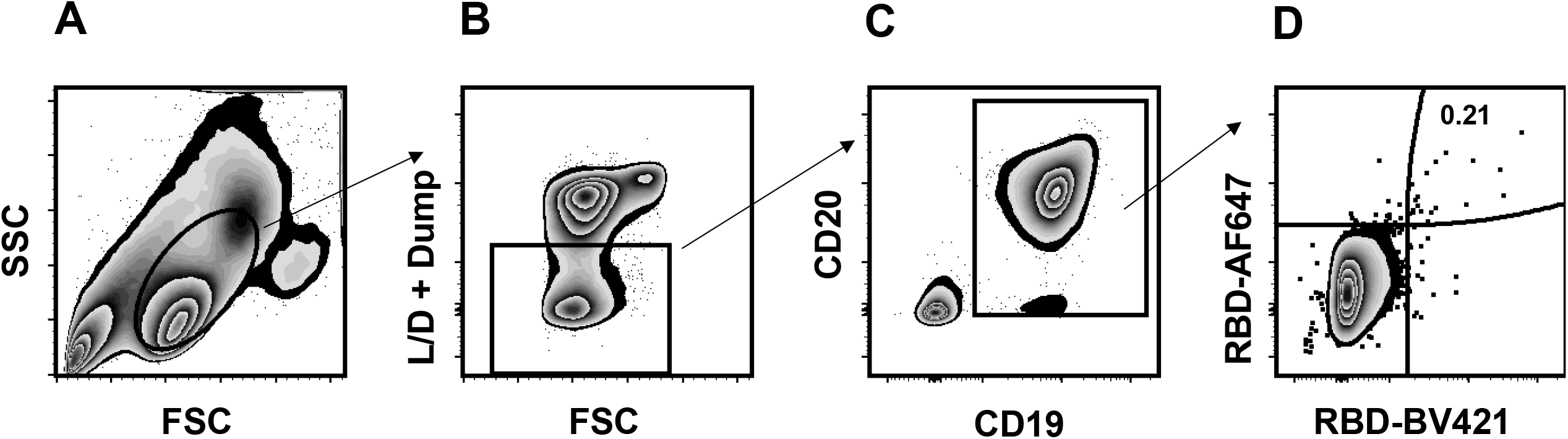
Gating strategy for SARS-CoV-2 S1 RBD-specific memory B cells. (**A**) Physical parameters; (**B**) Exclusion of dead cells and non-B cells (CD14^+^, CD3^+^, CD4^+^, CD16^+^); (**C**) CD19^+^CD20^Lo^ B cells were further gated to distinguish (**D**) RBD-specific B cells based on dual labeling in the same staining tube with two fluorescent RBD tetramers separately conjugated with Alexa Fluor 647 and BV421.

